# Osteoporosis Prevalence in Cardiovascular Kidney Metabolic Syndrome: Implications for Mortality

**DOI:** 10.1101/2025.02.10.25322036

**Authors:** Xi Du, Qianyu Yang, Zhiqiang Duan, Hui Gu, Xue Zhang, Hao Liu, Mao Xiao, Zhuoxing Li, Ming Zhao, Xiang Xiao

## Abstract

**Objective:** This study aims to investigate the prevalence of osteoporosis (OS) in patients with Cardiovascular Kidney Metabolic (CKM) Syndrome and its relationship with the risk of mortality in CKM patients.

**Methods:** Patients diagnosed with CKM were included from the National Health and Nutrition Examination Survey database between 2009 and 2018. The primary endpoint was all-cause mortality, while the secondary endpoint was cardiovascular mortality. The relationship between OS and the prognosis of all-cause mortality and cardiovascular mortality in CKM patients was assessed using Kaplan-Meier survival curves and multivariate Cox regression model.

**Results:** A total of 602 Non-CKM (stage 0) and 6129 CKM (stages 1, 2, 3, and 4) patients were included. The prevalence of OS in CKM patients at stages 0, 1, 2, 3, and 4 was 3.49%, 2.25%, 4.97%, 13.52%, and 8.39%, respectively. The multivariate Cox regression analysis revealed that compared to the normal bone mineral density group, CKM patients in the osteopenia group had a 55% increased risk of all-cause mortality (HR=1.55; 95% CI, 1.22-1.98, *P* = 0.001) and a 120% increase in the risk of cardiovascular mortality (HR=2.20; 95% CI, 1.39-3.46, *P* = 0.001); In the OS group, CKM patients exhibited a 199% increased risk of all-cause mortality (HR = 2.99; 95% CI, 1.86-4.82, *P* = 0.001) and a 2.14-fold increase in the risk of cardiovascular mortality (HR=3.14; 95% CI, 1.09-9.02, *P* = 0.03).

**Conclusion:** Osteopenia and OS are more prevalent in patients with CKM and are associated with an increased risk of all-cause and cardiovascular mortality.

## 1. Introduction

The cardiovascular-renal-metabolic (CKM) syndrome is a scientific statement proposed by the American Heart Association (AHA). The CKM syndrome is defined as a systemic disease characterized by the pathophysiological interactions between metabolic risk factors, chronic kidney disease (CKD), and the cardiovascular system, leading to a high incidence of multiple organ dysfunction and adverse cardiovascular outcomes ^[1]^. Furthermore, it reflects the interactions between metabolic risk factors, CKD, and the cardiovascular system, which have profound effects on morbidity and mortality ^[2]^.

The definition of osteoporosis (OS) is “a systemic skeletal disease characterized by a reduction in bone mass and microarchitectural deterioration of bone tissue, leading to an increased fragility of bones and a heightened risk of fractures” ^[3]^. It is not only a common disease in aging societies but also an important risk factor for various chronic diseases. Many risk factors associated with OS include low peak bone mass achieved during growth, hormonal factors, the use of certain medications, smoking, low physical activity, inadequate calcium and vitamin D intake, ethnicity, short stature, and a personal or family history of fractures ^[4]^. Moreover, the risk factors also include hyperlipidemia, hypertension, diabetes mellitus, oxidative stress, inflammation, Hyperhomocysteinemia, cardiovascular diseases, among others ^[5, 6]^. OS, as a metabolic bone disease^[7]^. It may also coexist with other metabolic diseases, including but not limited to obesity, diabetes, non-alcoholic fatty liver disease, dyslipidemia, and cardiovascular diseases (CVD) ^[8]^. Current research findings indicate that OS may be associated with various metabolic-related diseases, including CKD, hyperparathyroidism, and atherosclerosis ^[9–11]^. However, since the concept of CKM was put forward, there has been no research on its incidence and interrelationships in CKM yet.

CKM patients often exhibit various factors associated with the occurrence of OS, which may lead to a high prevalence of OS within this population; moreover, the condition of OS itself may further complicate the pathology and unfavorable prognosis of CKM patients. Therefore, elucidating the relationship between the two is essential for a more comprehensive understanding of the pathophysiological mechanisms of CKM. This understanding is critically important for reducing cardiovascular and all-cause mortality risks in CKM patients and improving their quality of life, and it provides a basis for developing more effective prevention and treatment strategies.

Thus, our study aims to explore the prevalence of OS among patients with CKM syndrome and its relationship with the risk of mortality in these patients.

## 2. Methods

### 2.1 Research Design and Participants

The data for this study is derived from the National Health and Nutrition Examination Survey (NHANES) database from 2009 to 2018, with CKM stage 0 defined as Non-CKM and CKM stages 1-4 defined as CKM. NHANES aims to explore individual-level demographic, health, and nutrition information through personal interviews and standardized physical examinations conducted by Mobile Examination Centers (MEC), and to assess the health and nutritional status of non-institutionalized civilians in the United States^[12]^.

In this study, in order to ensure the accuracy and credibility of the research, we adopted the staging structure of CKM syndrome proposed by the American Heart Association: 1) Stage 0, no CKM risk factors; 2) Stage 1, excessive or dysfunctional fat; 3) Stage 2, metabolic risk factors (hypertriglyceridemia, hypertension, diabetes, metabolic syndrome) or moderate to high-risk CKD; 4) Stage 3, subclinical cardiovascular disease or risk equivalents in CKM syndrome (high predicted cardiovascular disease risk or very high-risk CKD); 5) Stage 4, clinical cardiovascular disease in CKM syndrome^[2]^ (**Supplementar table 1)**. The diagnosis and classification of OS utilized dual-energy X-ray absorptiometry (DEXA)^[13]^. In the NHANES database, OS is defined as (1) a T-score of ≤ −2.5 for the femoral neck or lumbar spine; (2) occurrence of a low-trauma hip fracture (regardless of bone density), or clinical fractures of the vertebrae, proximal humerus, pelvis, or distal forearm with T-scores > −2.5 but < −1.0; or (3) a fracture risk assessment tool (FRAX) score that meets the intervention threshold set by the National Osteoporosis Foundation (hip fracture risk ≥ 3% or major osteoporotic fracture risk ≥ 20%), fulfilling any one of the above criteria^[14]^. Exclusion criteria: 1) Absence of CKM diagnosis, 2) Age under 20 years, 3) Pregnancy, 4) Missing osteoporosis-related data. All-cause mortality is defined as the termination of an individual’s life due to any cause, while cardiovascular death is defined as the death directly caused by diseases related to the cardiovascular system^[15]^. The definitions of smoking, drinking, hypertension, anemia, hyperlipidemia, diabetes,CKD, etc. can be found in **Supplementary Table 1**.

NHANES is a survey project conducted by the National Center for Health Statistics of the Centers for Disease Control and Prevention (CDC) and has been approved by the Institutional Review Board of the National Center for Health Statistics. The study protocol adheres to the ethical standards set forth in the Declaration of Helsinki of 1964 and its subsequent amendments, and has received approval from the National Health Statistics Research Ethics Review Board. All participants signed informed consent forms^[16]^.

### 2.2. Statistical analysis

According to the guidelines of the CDC in the United States, the statistical methods used in this study accounted for sample weights. Count data were described using means (standard error, SE), and categorical variables were described using percentages (SE). A multivariable Cox regression model was employed to evaluate the relationship between OS and the risk of all-cause mortality and cardiovascular mortality in CKM patients. Kaplan-Meier survival curves were used to assess the relationship between OS and the prognosis of all-cause mortality and cardiovascular mortality in CKM patients. The method of deletion is used to handle missing values. In the sensitivity analysis, multiple imputation is employed to address the missing values, in order to eliminate the bias caused by the deletion of missing data. All statistical analyses were performed using R version 4.3.1, and a two-sided *P* value of <0.05 was considered statistically significant.

## 3. Results

### 3.1 Baseline characteristics

Our study included data from 49,693 patients in the NHANES database from 2009 to 2018. After exclusions, a total of 602 Non-CKM (CKM stage 0) and 6129 CKM patients (CKM stage 1, 2, 3, 4) were finally included, among which the proportion of stage 1 patients was 26.36%, stage 2 patients 60.27%, stage 3 patients 3.74%, and stage 4 patients 9.62%. In CKM patients, the average age was 49.08 years, with males constituting 52.08% of the sample. The average BMI was 29.51 kg/m², and the incidence rates of hypertension, diabetes, CKD, and hyperlipidemia were 42.57%, 16.81%, 14.92%, and 72.11%, respectively. Compared with the normal bone mineral density (BMD) population, patients with osteopenia and OS were older, had lower eGFR and BMI, and had a higher proportion of females, as well as higher incidence rates of hypertension, hyperlipidemia, diabetes, and CKD (All *P* < 0.05) (**Figure 1**,**Table 1)**.

**Figure 1.**
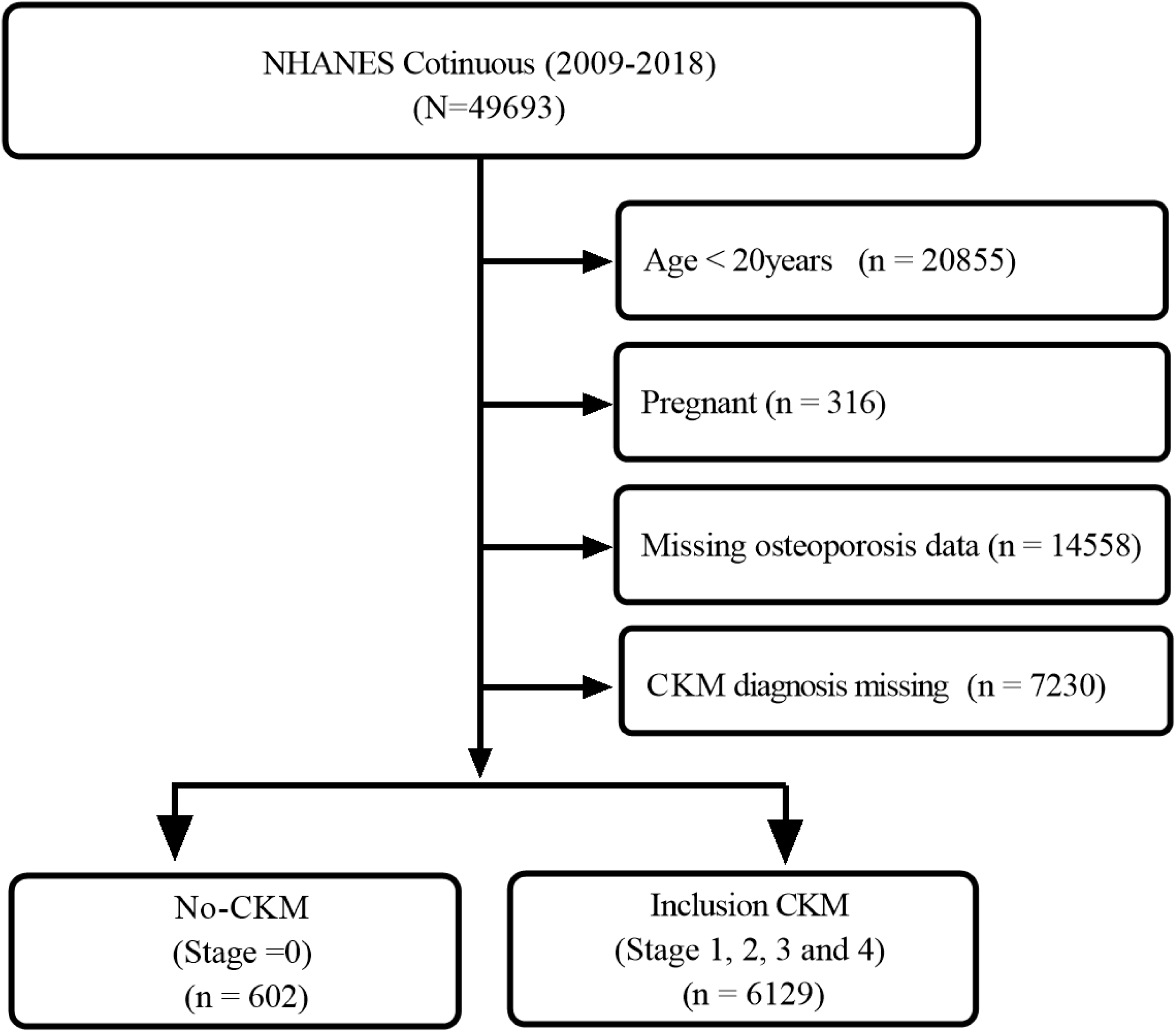
Inclusion flowchart.

**Table 1.**
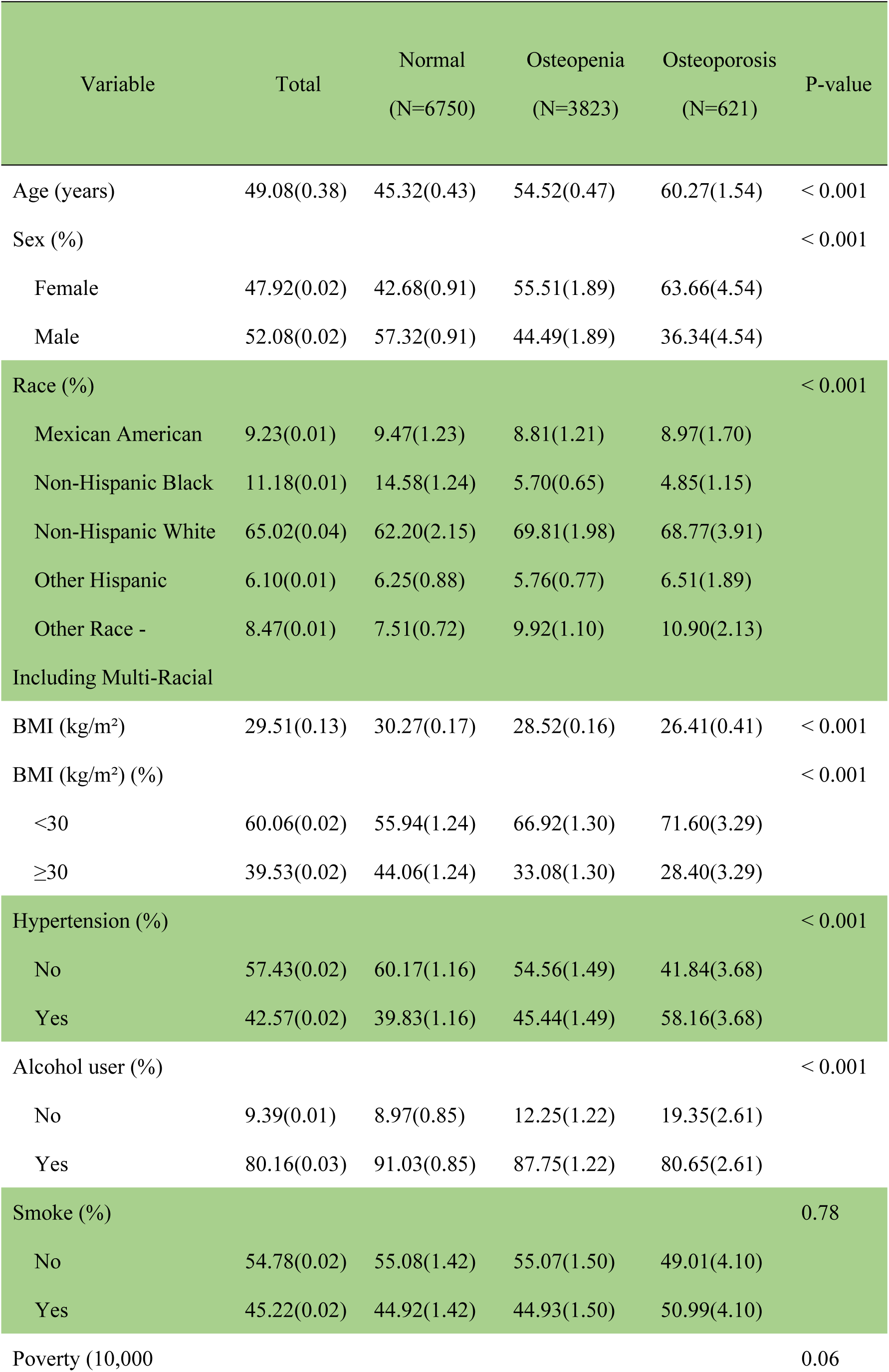
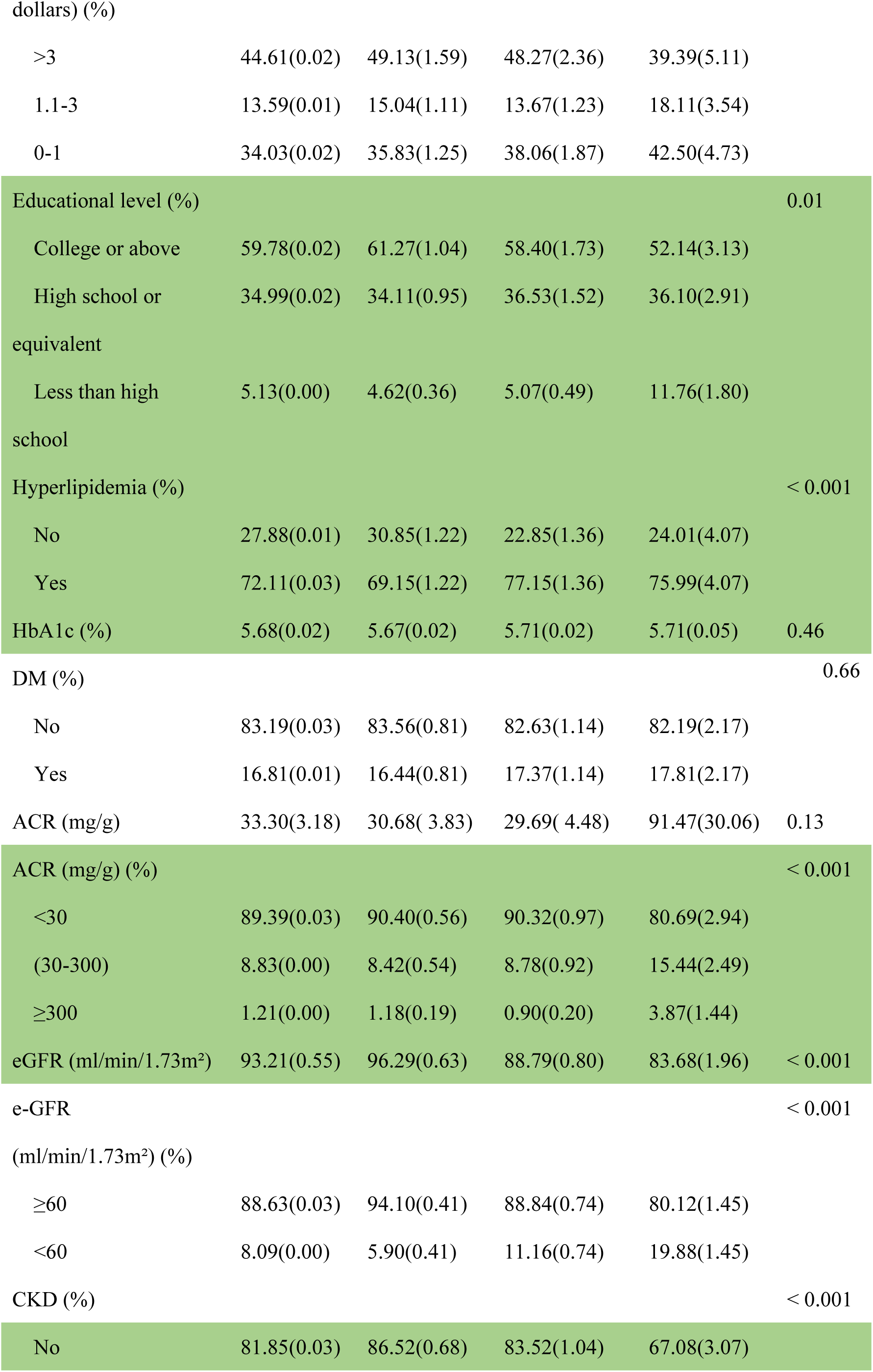
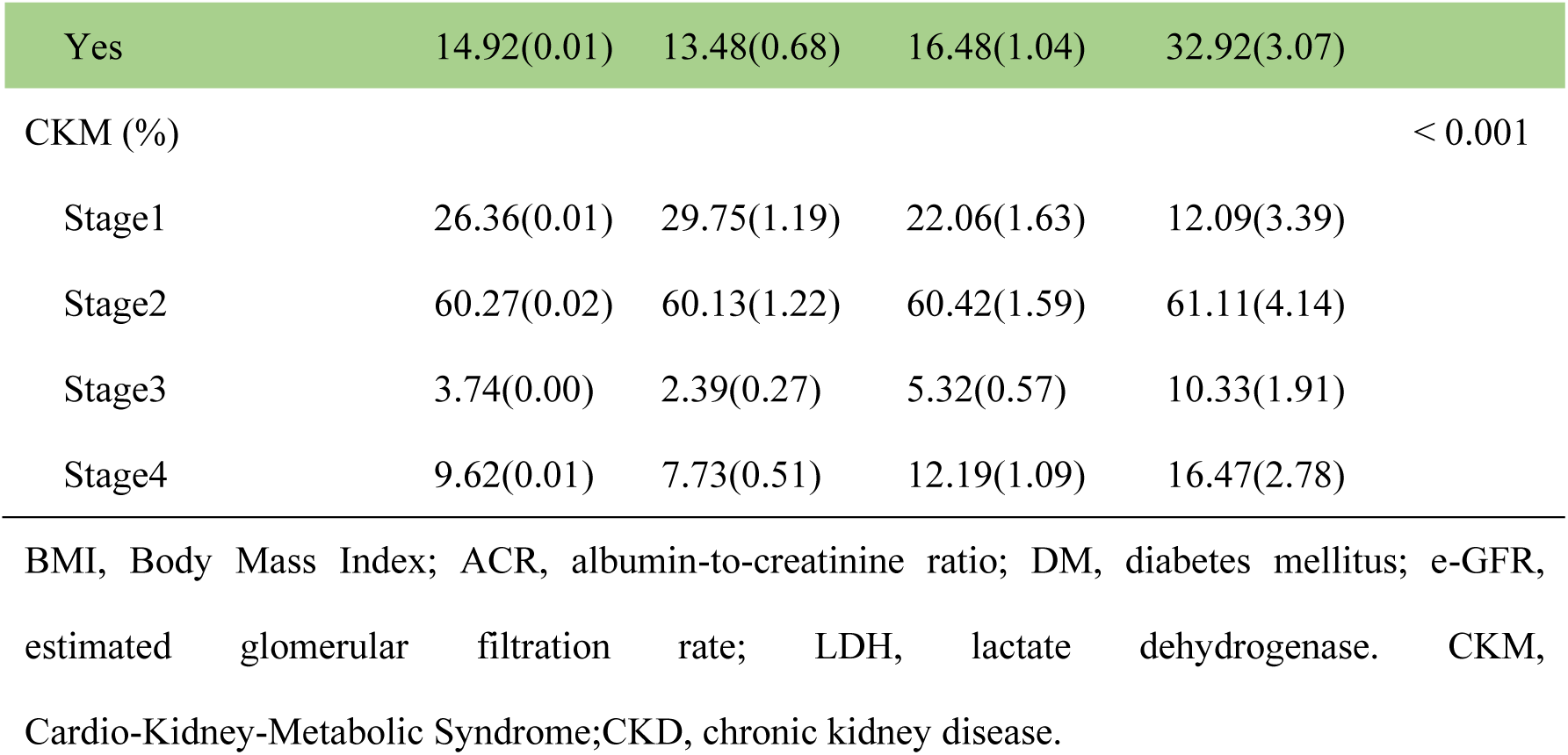
Baseline clinical features of enrolled individuals.

### 3.2 Incidence of OS in patients with CKM

In Non-CKM (stage 0) patients, the proportions of normal BMD, osteopenia, and OS were 63,56%, 32.95%, and 3.49%, respectively. However, among the 6129 CKM patients included, individuals with normal BMD accounted for 61.19%, those with osteopenia accounted for 33.70%, and those with OS accounted for 5.12%. Specifically, in CKM stage 1 patients, the proportions of normal BMD, osteopenia, and OS were 70.24%, 27.52%, and 2.25%, respectively; in CKM stage 2 patients, the proportions were 62.08%, 32.95%, and 4.97%; in CKM stage 3 patients, the proportions were 39.73%, 46.75%, and 13.52%; and in CKM stage 4 patients, the proportions were 49.98%, 41.64%, and 8.39% (**Supplementary table 2**). Among all CKM patients, the incidence of osteopenia in male was 28.08%, and that of OS was 3.42%, while in female, the incidence of osteopenia was 38.08%, and that of OS was 6.51%. The incidence rates of both osteopenia and OS in female CKM patients were significantly higher than those in male CKM patients (*P* <0.001) (**Supplementary table 2**). For age of CKM patients < 65 years, the proportions of osteopenia and OS were 28.68% and 3.68%, respectively, while for age of CKM patients ≥ 65 years, the proportions were 49.54% and 9.74%, indicating a higher risk of bone-related issues in elderly CKM patients (*P* < 0.001) (**Supplementary table 2**). Among CKM patients over 65 years of age, the incidence of osteopenia in female was 58.97%, and that of OS was 15,16%; for male, the incidence of osteopenia was 38.54%, and that of OS was 3.42%. The prevalence of both osteopenia and OS was higher in female (All *P* < 0.001) (**Supplementary table 2**). When investigating the prevalence of OS among CKM patients from different ethnicities, we found that the osteopenia and OS in non-Hispanic black individuals were significantly lower than in other ethnicities (All *P* < 0.001) (**Supplementary table 2**).

### 3.3 Primary outcome: risk of all-cause mortality

#### 3.3.1 Kaplan-Meier survival analysis

By performing Kaplan-Meier survival analysis, the relationship between bone status and the risk of all-cause mortality in CKM patients was assessed after adjusting for age (‘< 60’ years, ‘≥ 60’ years), gender (‘Female’, ‘Male’), race (‘Mexican American’, ‘Non-Hispanic Black’, ‘Non-Hispanic White’, ‘Other Hispanic’, ‘Other Race - Including Multi-Racial’), BMI (< 30kg/m^2^, ≥ 30 kg/m^2^), smoke (‘yes’ or ‘no’), alcohol use (‘yes’ or ‘no’), education (‘College or above’, ‘High school or equivalent’, ‘Less than high school’), poverty (‘0-1’, ‘1.1-3’, ‘> 3’), anemia (‘yes’ or ‘no’), hyperlipidemia (‘yes’ or ‘no’), hypertension (‘yes’ or ‘no’), DM (‘yes’ or ‘no’), RAASi use (‘yes’ or ‘no’) and other factors, the results indicate that patients with normal BMD group in the CKM had better survival outcomes compared to those in the osteopenia group, whereas patients in the OS group exhibited the poorest survival outcomes (Log-rank *P* < 0.001) (**Figure 2A)**.

**Figure 2.**
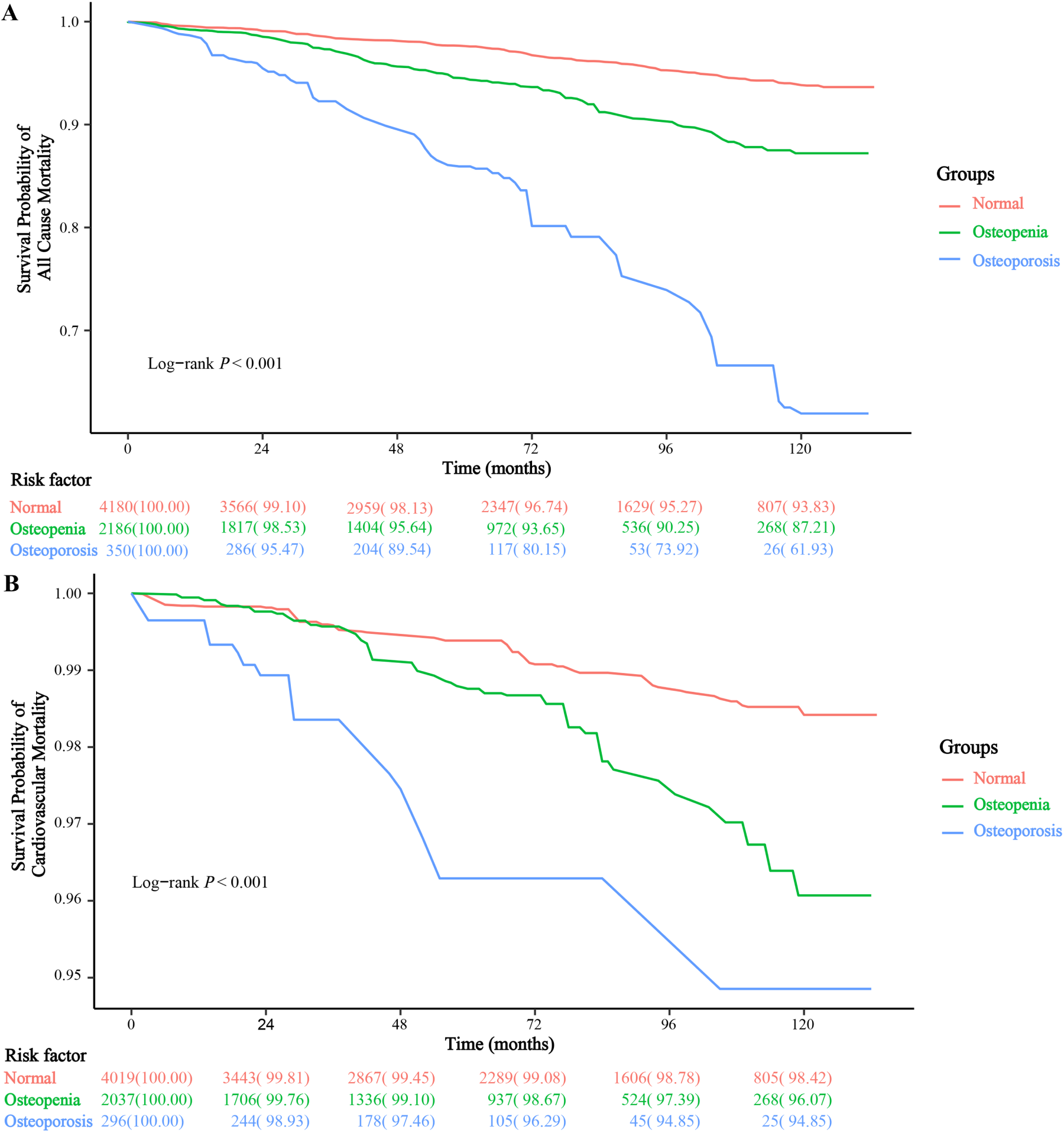
The relationship between osteoporosis and the prognosis of patients with CKM through Kaplan-Meier survival analysis. (A) all-cause mortality; (B) cardiovascular mortality. CKM, Cardiovascular-kidney-Metabolic.

#### 3.3.2 Multivariate Cox regression analysis

Through a multifactorial Cox regression model, after adjusting for multiple variables, the results indicated that, compared to the normal BMD group, CKM patients with osteopenia had a 55% increased risk of all-cause mortality (HR = 1.55; 95% CI, 1.22-1.98, *P* = 0.001), while CKM patients with OS exhibited a 1.99-fold increase in all-cause mortality risk (HR = 2.99; 95% CI, 1.86-4.82, *P* = 0.001) (**Supplementary -table 3, Figure 3**).

**Figure 3.**
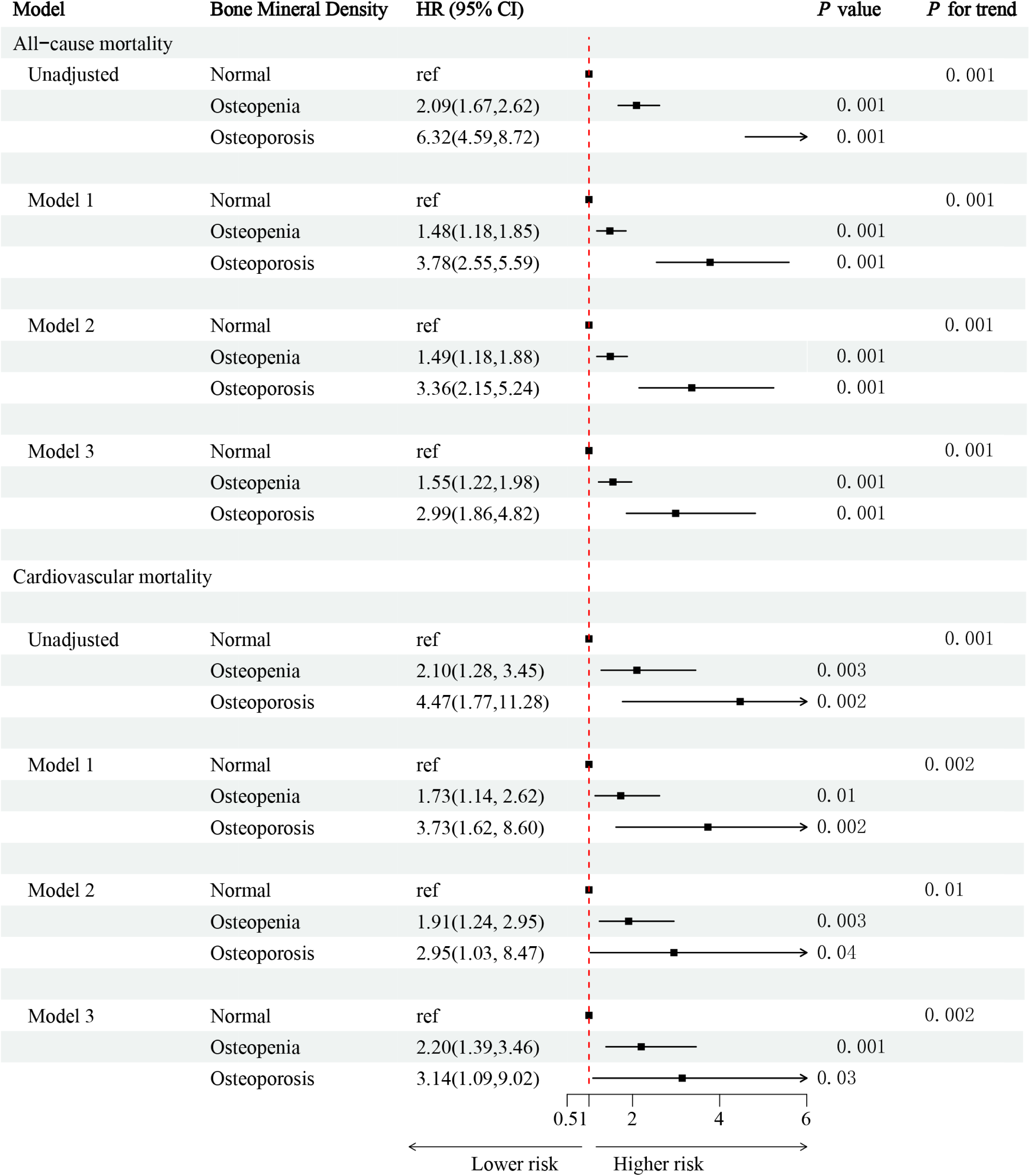
Association between osteoporosis and all-cause mortality. **Model 1** adjusted for baseline age (‘<60’ years, ‘≥60’ years), gender (‘Female’, ‘Male’), race (‘Mexican American’, ‘Non-Hispanic Black’, ‘Non-Hispanic White’, ‘Other Hispanic’, ‘Other Race - Including Multi-Racial’), BMI (<30kg/m^2^, ≥30 kg/m^2^); **Model 2** adjusted for covariates in model 1 plus smoke (‘yes’ or ‘no’), alcohol use (‘yes’ or ‘no’), education (‘College or above’, ‘High school or equivalent’, ‘Less than high school’), poverty (‘0-1’, ‘1.1-3’, ‘>3’). **Model 3** adjusted for covariates in model 2 plus hyperlipidemia (‘yes’ or ‘no’), hypertension (‘yes’ or ‘no’), DM (‘yes’ or ‘no’). HR, Hazard ratio; CI, Confidence interval; BMI, Body Mass Index; RAASi, renin-angiotensin system inhibitors; CKD, chronic kidney disease; DM, diabetes mellitus; CKM, Cardiovascular-kidney-Metabolic.

**Figure 4.** Association between osteoporosis and cardiovascular mortality. **Model 1** adjusted for baseline age (‘<60’ years, ‘≥60’ years), gender (‘Female’, ‘Male’), race (‘Mexican American’, ‘Non-Hispanic Black’, ‘Non-Hispanic White’, ‘Other Hispanic’, ‘Other Race - Including Multi-Racial’), BMI (<30kg/m^2^, ≥30 kg/m^2^); **Model 2** adjusted for covariates in model 1 plus smoke (‘yes’ or ‘no’), alcohol use (‘yes’ or ‘no’), education (‘College or above’, ‘High school or equivalent’, ‘Less than high school’), poverty (‘0-1’, ‘1.1-3’, ‘>3’). **Model 3** adjusted for covariates in model 2 plus hyperlipidemia (‘yes’ or ‘no’), hypertension (‘yes’ or ‘no’), DM (‘yes’ or ‘no’). HR, Hazard ratio; CI, Confidence interval; BMI, Body Mass Index; RAASi, renin-angiotensin system inhibitors; CKD, chronic kidney disease; DM, diabetes mellitus; CKM, Cardiovascular-kidney-Metabolic.

#### 3.3.3 Stratified analysis

In stratified analysis, we assessed whether the relationship between all-cause mortality risk in CKM patients was influenced by factors such as age (’<60’ years, ‘≥ 60’ years), gender (’Female’, ‘Male’), race (’Mexican American’, ‘Non-Hispanic Black’, ‘Non-Hispanic White’, ‘Other Hispanic’, ‘Other Race - Including Multi-Racial’), BMI (< 30 kg/m², ≥ 30 kg/m²), hypertension (’yes’ or ‘no’), diabetes (’yes’ or ‘no’), CKD (’yes’ or ‘no’), and stages of CKM (Stage 1, 2, 3, and 4). The results indicated that, except for age (*P* for interaction < 0.05), there was no interaction between OS and variables such as gender, race, BMI, hypertension, diabetes, CKD, and stages of CKM (*P* for interaction > 0.05) (**Table 2**).

**Table 2.**
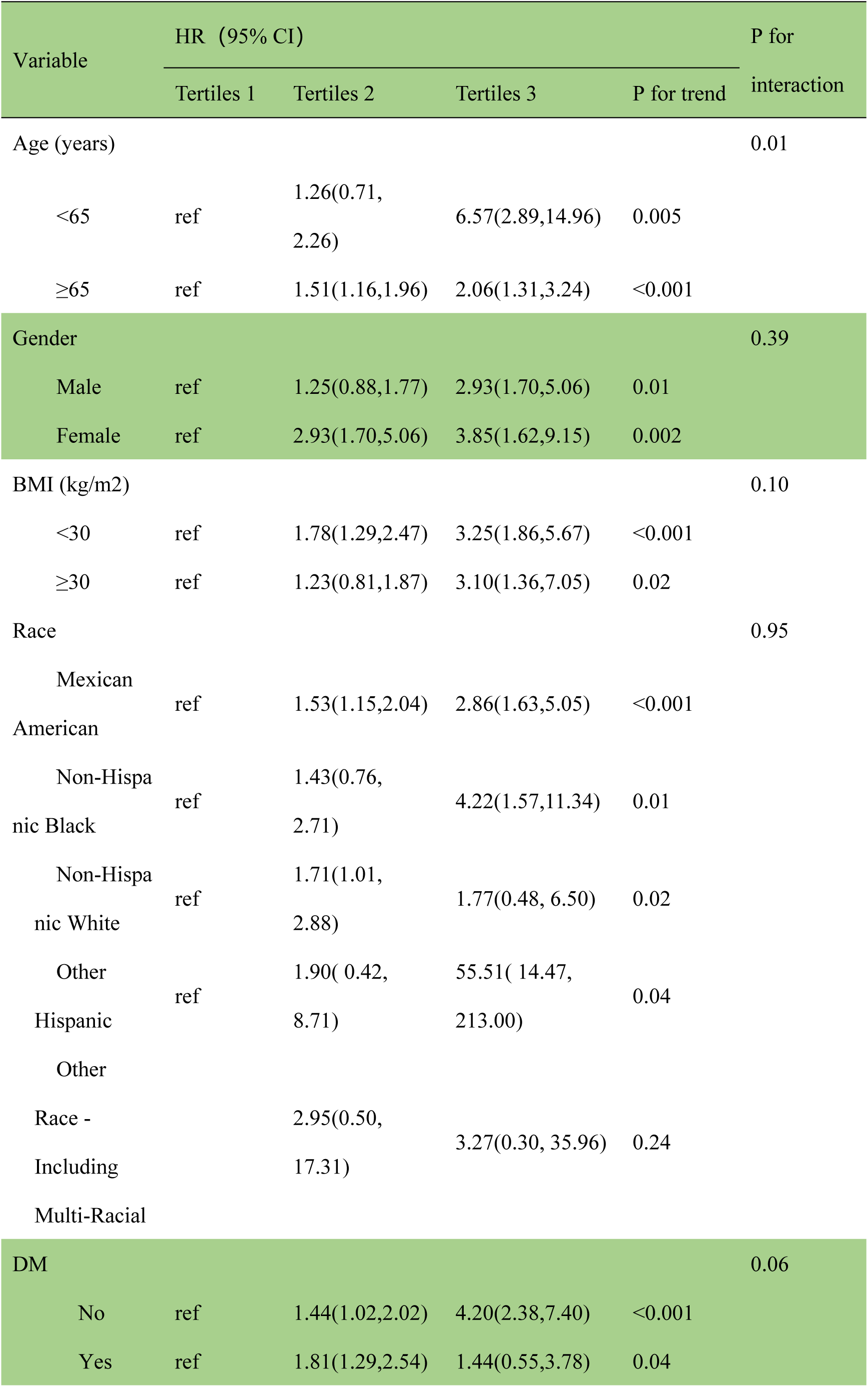
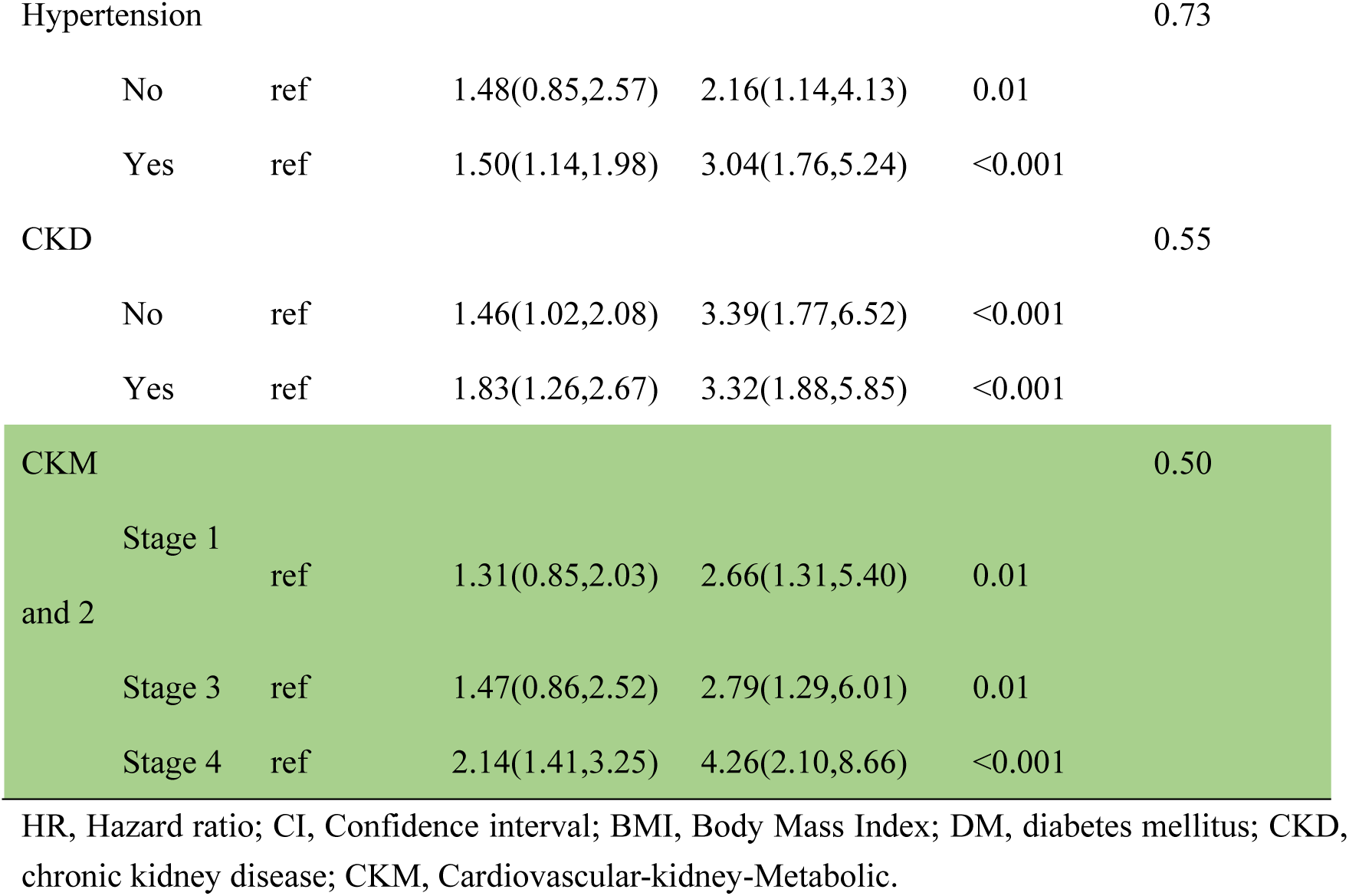
Stratified analysis of osteoporosis and the risk of CKM in individuals.

#### 3.3.4 Sensitivity analysis

In order to mitigate the impact of missing values on the results, we performed multiple imputation on the data and re-evaluated the relationship between OS and the risk of all-cause mortality in patients with CKM. After multivariable adjustment, the results of the Cox regression analysis indicated that, using individuals with normal BMD as the reference, the risk of all-cause mortality in the osteopenia group was 1.55 times that of the normal BMD group (HR = 1.55; 95% CI, 1.22-1.98, *P* < 0.001), while the risk of all-cause mortality in the OS group was 1.99 times that of the normal BMD group (HR = 2.99; 95% CI, 1.86-4.82, *P* < 0.001) (**Supplementary table 4**).

### 3.4 Secondary outcome: cardiovascular mortality

#### 3.4.1 The KM survival analysis evaluates the relationship between OS and cardiovascular mortality prognosis in CKM patients

By performing Kaplan-Meier survival analysis to assess the relationship between bone status and the risk of cardiovascular mortality in CKM patients, after adjusting for factors such as age (’< 60’ years, ‘≥ 60’ years), gender (’Female’, ‘Male’), race (’Mexican American’, ‘Non-Hispanic Black’, ‘Non-Hispanic White’, ‘Other Hispanic’, ‘Other Race - Including Multi-Racial’), BMI (< 30 kg/m², ≥ 30 kg/m²), smoking (’yes’ or ‘no’), alcohol use (’yes’ or ‘no’), education (’College or above’, ‘High school or equivalent’, ‘Less than high school’), poverty (’0-1’, ‘1.1-3’, ‘> 3’), anemia (’yes’ or ‘no’), hyperlipidemia (’yes’ or ‘no’), hypertension (’yes’ or ‘no’), diabetes (’yes’ or ‘no’), and RAAS inhibitor use (’yes’ or ‘no’), the results showed that CKM patients in the normal BMD group had better survival than those in the osteopenia group, while patients in the OS group had the worst survival outcomes (Log-rank *P* < 0.001) (**Figure 2B**).

#### 3.4.2 Multivariate Cox regression analysis

Using a multivariate Cox regression model to evaluate the relationship between OS and cardiovascular mortality risk in CKM patients, the results showed that after adjusting for multiple variables, compared to the normal BMD group, the cardiovascular mortality risk in CKM patients with osteopenia increased by 120% (HR = 2.20; 95% CI, 1.39-3.46, *P* < 0.001), while patients with OS had a 2.14-fold increase in cardiovascular mortality risk (HR = 3.14; 95% CI, 1.09-9.02, *P* = 0.03) (**Supplementary - table 5**, **Figure 3**).

#### 3.4.3 Sensitivity analysis

Multiple imputation was performed on the data, and the relationship between OS and the risk of cardiovascular mortality in patients with CKM was reassessed. After multivariable adjustment, the results of the Cox regression analysis showed that, taking the group with normal BMD as the reference, the risk of cardiovascular mortality in the osteopenia group was 1.20 times higher than that of the normal BMD group (HR = 2.20; 95% CI, 1.39-3.46, *P* < 0.001), and the risk of cardiovascular mortality in the OS group was 2.14 times higher than that of the normal BMD group (HR = 3.14; 95% CI, 1.09-9.02, *P* = 0.03) (**Supplementary table 6**).

## 4. Discussion

Our study findings indicate that among the 602 Non-CKM individual and 6129 patients with CKM included in the study, the proportion of individuals with osteopenia is 32.87%, while the proportion with OS is 4.90%. Compared to non-CKM individuals, patients with CKM exhibit a higher incidence of osteopenia and OS, particularly those in CKM stage 3, which is the most pronounced. Furthermore, the occurrence of either osteopenia or OS significantly increases the risk of all-cause and cardiovascular mortality in CKM patients.

OS is defined as a systemic skeletal disease characterized by a reduction in bone mass and alterations in the microstructure of bone tissue, leading to increased bone fragility and an elevated risk of fractures ^[3]^. The World Health Organization (WHO) defines OS based on BMD measurements obtained through DXA, which is currently employed worldwide ^[17]^. Salari N et al. ^[18]^ conducted a systematic review and meta-analysis of 86 studies according to PRISMA standards, revealing that the global prevalence of OS stands at 18.3%, with a prevalence of 23.1% in women and 11.7% in men worldwide. Xiao PL et al. ^[19]^ estimated the global and regional prevalence of OS by analyzing 57,933 citations, 108 individual studies, and 343,704 participants. Their results indicated significant variability in prevalence rates among countries (ranging from 4.1% in the Netherlands to 52.0% in Turkey) and continents (ranging from 8.0% in Oceania to 26.9% in Africa). As of 2022, the global prevalence of OS and osteopenia was estimated to be 19.7% and 40.4%, respectively. In the cohort of CKM patients from the United States that we included, the proportion of individuals with osteopenia is 32.87%, while the proportion with OS is 4.90%. However, compared to Non-CKM patients of the same period, the proportions of osteopenia and OS are 32.95% and 3.49%, respectively. Overall, compared to the prevalence rates reported in most literature, the prevalence of OS in the population included in NHANES from 2009 to 2018 shows a relatively lower level. In addition, the prevalence of osteopenia in CKM stages 0, 1, 2, 3, and 4 was 32.95%, 27.52%, 32.95%, 46.75%, and 41.64% respectively, while the prevalence of OS in CKM stages 0, 1, 2, 3, and 4 was 3.49%, 2.25%, 4.97%, 13.52%, and 8.39% respectively. Therefore, we can observe that during CKM stages 1 the prevalence of osteopenia and OS is lower compared to that of the healthy population in the United States; however, in CKM stage 2, 3 and 4, the prevalence of osteopenia and OS significantly increases compared to the Non-CKM (CKM stages 0). The overall osteopenia and OS prevalence were the highest in patients with stage CKM stage 3. However, we also observe that although CKM patients have a higher prevalence of OS, CKM stage 1 patients have a lower prevalence of OS compared to Non-CKM individuals. This suggests that patients with weight gain may have relatively higher intake of calcium or vitamin D, which helps reduce the risk of OS prevalence. However, as the disease progresses, this benefit might be offset by factors such as the gradual onset of CKD, DM, or CVD in the patients, resulting in higher OS prevalence in CKM stages 2, 3, and 4. Furthermore, we find that in CKM stage 4 patients, the prevalence of OS decreases compared to CKM stage 3, which may be due to increased attention to these individuals and a higher occurrence of mortality.

However, why is the prevalence of OS significantly increased in CKM patients? Its possibly due to multiple functional disturbances in the cardiovascular, renal, and metabolic systems, with the pathophysiological changes likely promoting the formation of OS. In terms of metabolism, the abnormal glucose metabolism and dyslipidemia in CKM patients may contribute to the risk of OS^[20]^. Hyperglycemia is exacerbated in diabetes and aging, leading to the excessive accumulation of advanced glycation end products (AGEs/AGOEs) within the bone. These products disrupt bone remodeling and reduce bone quality by altering the organic matrix, mineral content, and water content of the bone, ultimately resulting in OS^[21]^. The role of osteoblasts is crucial for investigating the regulatory mechanisms of various types of OS, as they are derived from bone marrow mesenchymal stem cells (BM-MSCs)^[22, 23]^. BM-MSCs are multipotent stem cells with the ability to differentiate into multiple lineages, serving as precursor cells for osteoblasts and adipocytes. Adipogenic differentiation represents another primary differentiation pathway that balances with osteogenic activity^[24]^. Therefore, changes in lipid metabolism in the body may trigger alterations in bone metabolism. Dysregulation of lipid metabolism in certain disease states can result in a cascade of effects on bone metabolism, ultimately leading to OS. In terms of cardiovascular implications, abnormal vascular function and hemodynamic changes in CKM patients may promote the development of OS. Blood vessels play a crucial role in bone homeostasis and repair; compromised blood flow dynamics can lead to deficiencies in angiogenesis and osteogenesis^[25]^, thus accelerating the progression of OS. In terms of the kidneys, patients with CKD due to CKM may experience renal osteodystrophy (ROD) as a result of comorbid CKD, which subsequently promotes the development of OS^[26]^. Furthermore, patients with CKD may experience abnormalities in calcium, phosphate, parathyroid hormone (PTH), and vitamin D due to CKD-mineral and bone disorder (CKD-MBD), resulting in abnormalities in bone turnover, mineralization, volume, linear growth, or strength. These changes can promote the occurrence of OS ^[27]^. However, with the increase in CKM stages, on the basis of metabolic abnormalities, the prevalence of cardio - renal - related diseases in patients also increases significantly. This may lead to a lower prevalence of osteopenia and osteoporosis in CKM stage 1 compared to CKM stage 0. However, in CKM stages 2 - 4, the prevalence of osteopenia and osteoporosis increases significantly. Therefore, this result also indicates that we particularly need to strengthen the screening related to bone health in patients with CKM stages 2 - 4. Therefore, we speculate that in CKM stages 1, the intake of proteins, calcium, and vitamin D by patients may counterbalance the bone changes aggravated by metabolic factors. However, in CKM stage 2-4, such intake is insufficient to compensate for the bone damage caused by metabolic factors, CKD and diabetes.

OS significantly increases the risk of mortality. Current studies have shown that untreated OS can lead to a vicious cycle of recurrent fractures, which typically results in disability and premature death^[28]^ ^[29]^. Data from observational studies have reported that osteoporotic fractures are associated with an increased risk of mortality, and the incidence of osteoporotic fractures increases exponentially in later life, paralleling the progression of frailty and the associated risk of mortality^[30]^. Our research results further indicate that in patients with CKM, the occurrence of OS and bone loss significantly increases the risks of all-cause mortality and cardiovascular mortality. Various comorbidities may also be potential factors influencing the association between the exacerbation of OS and the elevated mortality risk in CKM patients. Chen X et al.^[31]^ found that the coexistence of a history of fractures and hypertension increases the all-cause mortality risk associated with OS. Li W et al.^[32]^ analyzed 18,658 subjects from the National Health and Nutrition Examination Survey and discovered an additive effect between diabetes and OS, which may exert a synergistic effect on all-cause mortality. Patients suffering from both conditions exhibit a higher risk of death, indicating that the onset of OS increases the mortality risk for diabetic patients.The study conducted by Nakano Y et al^[33]^ shows that OS is closely related to the decline in kidney function, which is associated with adverse survival outcomes and increased mortality risk in patients with CKD. Baldini V et al.^[5]^ found that the incidence of cardiovascular disease and cardiovascular mortality is related to a decrease in BMD and fractures.This may be due to the limited mobility of patients after fractures and increased bed rest time, which can subsequently lead to a series of complications such as pulmonary infections ^[34]^ and thrombosis ^[35]^. Therefore, various comorbidities, such as the coexistence of a history of fractures with hypertension, the synergistic effects of diabetes and OS, and the associations of OS with declining renal function and cardiovascular diseases, are likely to be potential factors behind the increased mortality risk in CKM patients following the onset of OS. This further highlights the importance of comprehensive management of comorbidities in CKM patients, which may contribute to improving patient prognosis. Additionally, in the stratified analysis, we found that abnormal bone mass can increase the risk of all-cause and cardiovascular mortality in CKM patients across different CKM stages. However, we also observed that this relationship exhibits heterogeneity across different age stratifications, being more prominent in elderly patients. This may be related to the higher incidence of OS in older patients, the presence of more comorbidities, and their reduced resilience to stressors.

Based on the results of this study, it is necessary to optimize current clinical treatment strategies to effectively reduce the mortality risk of CKM patients. Firstly, the presence of multiple comorbidities and their interactions must be fully considered during the treatment process. For CKM patients who also suffer from comorbidities such as hypertension, diabetes, and cardiovascular diseases, a comprehensive treatment plan should be devised to manage various conditions in a coordinated manner^[1, 36]^. Secondly, regarding OS as a critical factor, in addition to the aforementioned screening and intervention measures, it is essential to strengthen health education for patients to enhance their awareness of the dangers posed by OS. This will encourage them to actively cooperate with treatment and take proactive preventive measures, such as maintaining moderate exercise and avoiding falls, to reduce the likelihood of fractures. In particular, the screening frequency for patients with CKM stage 2-4 needs to be increased, along with a heightened focus on their condition. Finally, an effective patient follow-up mechanism should be established to closely monitor changes in patients’ conditions, timely adjust treatment regimens, and ensure the efficacy and sustainability of treatment, thereby minimizing the mortality risk for CKM patients to the greatest extent possible.

Although this study has made efforts to explore the related topics, it still has certain limitations. Firstly, from the perspective of the sample, the relatively small sample size poses challenges to the precision and representativeness of the results, making it difficult to comprehensively reflect the overall characteristics. Furthermore, during the research process, some potential confounding factors, such as details of patients’ lifestyles (including exercise habits, dietary structure, etc.) and various medication use, may not have been fully controlled, thereby affecting the results to some extent. Lastly, the inability to comprehensively assess CKM diagnostic-related indicators in the NHANSE may introduce some bias in the morbidity rates. To overcome the design flaws of the study, subsequent research could adopt a longitudinal design to conduct long-term follow-up observations of the subjects to clearly determine the causal relationships among the variables.

In summary, CKM has emerged as a syndrome for evaluating the heart, kidneys, and metabolic status, with its incidence and importance increasingly highlighted. OS has a high incidence among CKM patients, particularly in those with CKM stage 3-4. Furthermore, OS is closely associated with the risks of cardiovascular events and all-cause mortality, a relationship that does not change with the CKM stages. Clinical decision-makers need to provide different monitoring frequencies for bone metabolism status according to the various CKM stages, with the aim of formulating comprehensive intervention programs to reduce the risk of mortality. Although this study has certain limitations, it provides important insights and directions for further research.

## Funding

The Science and technology fund of Chengdu Medical College (CYZYB22-02).

The research fund of Sichuan Medical and Health Care Promotion institute (KY2022QN0309).

Sichuan Provincial Medical Association Youth Innovation Project (Q23021).

Health Commission of Sichuan Province Medical Science and Technology Program (24QNMP100).

Sichuan Provincial Geriatric Clinical Medicine Research Center (24LHLNYX1-48).

## Author Contributions

Conception and design of the study: Xi Du, Qianyu Yang, Zhiqiang Duan, Ming Zhao, Xiang Xiao;

Acquisition and analysis of data: Xi Du, Qianyu Yang, Zhiqiang Duan;

Drafting the manuscript or figures: Xi Du, Qianyu Yang, Zhiqiang Duan, Ming Zhao, Xiang Xiao.

## Data Availability

Some or all datasets generated or analyzed during the current study are not publicly available. However, they can be obtained from the corresponding author upon reasonable request.

## Acknowledgments

The authors express their gratitude to all participants of this study for their significant contributions.

## Ethical Approval

The study protocol adhered to the ethical standards set forth in the 1964 Declaration of Helsinki and its subsequent amendments. Approval was obtained from the National Committee for Ethical Review of Health Statistics Research, and all participants provided signed informed consent.

## Declarations

The authors state that there are no known financial conflicts of interest or personal relationships that could have influenced the research presented in this paper.

